# Pragmatic RE-AIM evaluation of the Train-the-Trainer Vaccine Champions Program in Vietnam

**DOI:** 10.64898/2025.12.16.25342229

**Authors:** Isabella Overmars, Jessica Kaufman, Thu-Anh Nguyen, Luong Tran, Thi Mai Nguyenc, Ngoc Anh Vu Nguyen, Chu Huu Trang, Suzanna Vidmar, Maharajan Muthu, Gregory Fox, Shiva Shrestha, Holly Seale, Margie Danchin

**Author notes:** Joint first authors (Isabella Overmars, Jessica Kaufman).

## Abstract

**Background and objective:** Vietnam had increasing rates of zero-dose children and sub-optimal COVID-19 vaccine booster coverage in 2022. With the Vietnamese Ministry of Health, we co-designed, implemented and evaluated a vaccine education and communication program for health workers and community leaders to improve trust, knowledge, communication skills and intention to vaccinate for routine childhood and COVID-19 vaccines.

**Method:** The Train-the-Trainer Vaccine Champions program was piloted in 2022 and deployed in 2023 in three low-coverage provinces. Master Trainers trained Provincial Trainers (healthcare workers and communication experts), who trained community leaders and other health workers as Champions to deliver community vaccine education sessions. Using mixed methods and the RE-AIM framework, we evaluated reach, effectiveness, adoption, implementation, and maintenance. Primary effectiveness outcomes included trust in vaccines and the healthcare system, vaccine knowledge, communication self-efficacy, satisfaction, and community respondents’ intention to vaccinate. Secondary outcomes included changes in Provincial level coverage pre- and 6-months post-intervention.

**Results:** The program trained 56 Provincial Trainers (47% female), 286 Vaccine Champions (46% female), and reached 4027 community attendees. Communication self-efficacy and trust in vaccines and the health system increased significantly among Provincial Trainers and Vaccine Champions post-training but change in knowledge was minimal. Participants reported high satisfaction with sessions and materials and 86% of community respondents reported increased intention to vaccinate. Champions continue to run education sessions and promote other health programs with the Ministry of Health.

**Conclusions:** In Vietnam, a multi-level Vaccine Champions program that trained healthcare workers and community leaders to promote routine childhood and COVID-19 vaccination increased participant trust in vaccines and the health system, self-efficacy, and community intention to vaccinate. The program has potential to reduce healthcare workforce burden and has ongoing government support. Hybrid implementation-effectiveness trials are needed to determine impact on vaccine coverage and cost-effectiveness, alongside implementation outcomes, to guide scalability.

## Introduction

Vaccination is critical to achieving sustainable development goal three (SDG3), ‘good health and wellbeing’.^1^ During the pandemic, achieving high coronavirus disease (COVID-19) vaccine acceptance and uptake was a priority, with a focus on vaccine equity and priority populations. At the same time, the pandemic precipitated unprecedented backsliding in childhood vaccine coverage.^2^ Despite early improvement in 2022, recovery has been slow. Zero-dose children, who have received no vaccines, increased from 13.9 million in 2022 to 14.5 million in 2023,^3^ and immunisation-related indicators for SDG3 are not on track to achieve the Immunisation Agenda targets of 90% coverage and no more than 6.5 million zero-dose children globally by 2030.^4^

Vietnam is among the top 20 countries globally with the most zero-dose children, with 187,315 under 12 months not receiving any vaccines in 2021.^2^ In 2022, the nation was also grappling with COVID-19 adult booster coverage that was below government targets, despite achieving 100% primary dose coverage in 2022.^5^ Many factors contributed to declines in childhood vaccine coverage in Vietnam including disruption to supply and health service delivery, and mandates and misinformation leading to decreased vaccine confidence, trust and uptake.^6, 7^

As the most trusted source of vaccine information, healthcare workers are the backbone of immunisation programs worldwide, and their recommendations are among the strongest predictors of vaccine uptake.^7–10^ However, being overburdened with responsibilities has led to burnout and staff shortages.^9, 11, 12^ In response to this issue, particularly during the pandemic, programs have endeavoured to engage diverse non-health community leaders to promote vaccines, especially amongst hard-to-reach and priority populations. While effects on uptake are varied,^13–16^ there is growing evidence for the role of well-trained community leaders in increasing trust in vaccines and the health system and increasing vaccine acceptance among their communities.^17–19^

Supported by the Australian Government and requested by the Vietnamese Ministry of Health, academics from Australia and Vietnam partnered with the United Nations Children’s Fund (UNICEF) to increase trust, knowledge, communication skills and intention to vaccinate for routine childhood and COVID-19 vaccines through a Train-the-Trainer Vaccine Champion education and communication skills program. This program aimed to upskill health and non-health workers to advocate for vaccines in three provinces, based on smaller programs delivered in Australia and Fiji,^18, 20^ and was evaluated using the RE-AIM implementation science framework.^21^

## Method

### Study design

Mixed methods were used to evaluate implementation of the Vaccine Champions program including the reach, effectiveness, adoption, implementation, and maintenance (RE-AIM).^21, 22^ Data were collected through quantitative and qualitative methods (Table 1), and reported using the Strengthening the Reporting of Observational Studies in Epidemiology (STROBE) checklist.^23, 24^

**Table 1:**
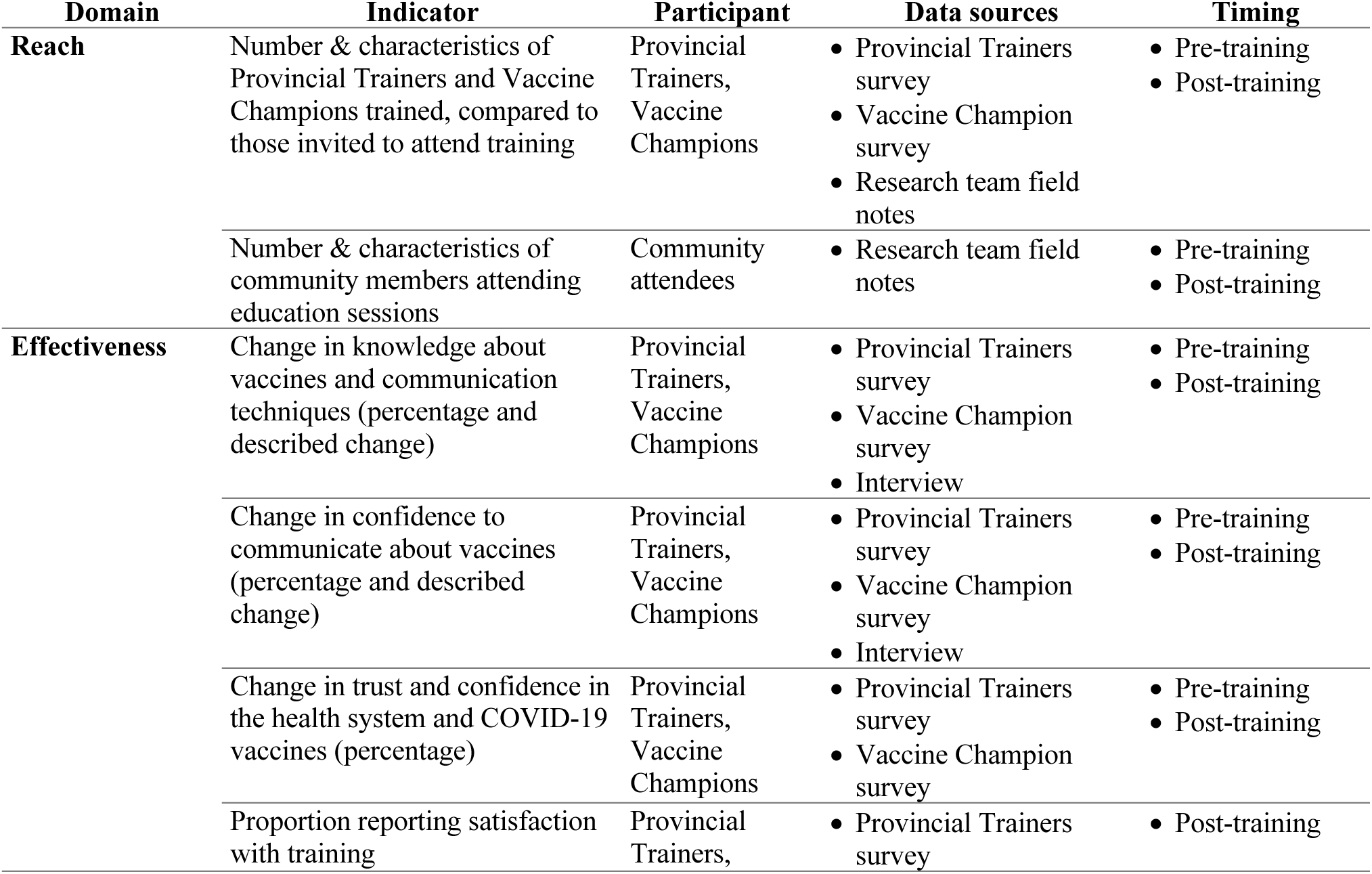
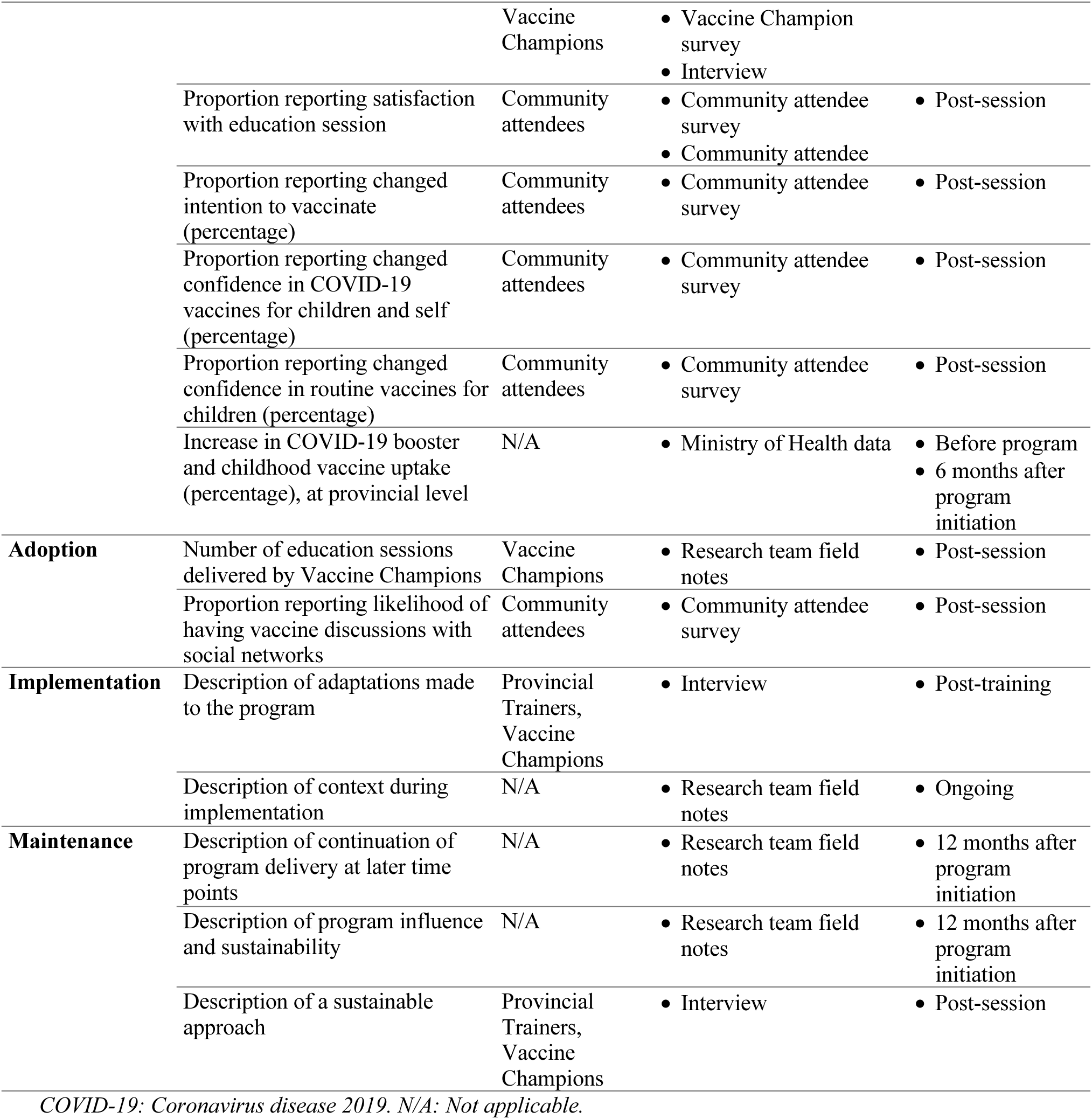
Data collection methods according to RE-AIM indicators.

### Setting

The program was implemented and evaluated in three of 58 provinces in Vietnam: Sóc Trăng, Gia Lai and Điện Biên. The Vietnamese Ministry of Health and UNICEF selected provinces with low COVID-19 adult booster and childhood vaccine coverage in 2022. Sóc Trăng is a coastal province in southern Vietnam, and home to ethnic minorities including Kinh, Khmer and Vietnamese Chinese. Gia Lai is mountainous, in the central highlands, and has ethnic groups including Gia Rai, Kinh and Bahnar. Điện Biên is mountainous, in the northwest, shares borders with Laos and China, and its primary ethnic groups are Thai and H’Mong. Downstream activities, referring to trainings and sessions run by trainees, were also undertaken in neighbouring provinces Đắk Lắk, Đăk Nông and Kon Tum. Data were collected between 1 April 2023 and 1 April 2024. Activities from 1 April 2024 until 31 December 2024 are reported under ‘Maintenance’.

### The Vaccine Champions Program

The Vietnamese Ministry of Health requested assistance from the Australian Expert Technical Assistance Program, co-ordinated by the Australian Department of Foreign Affairs and Trade, to improve COVID-19 and routine childhood vaccine acceptance and intention to vaccinate in 2022. The program’s theory of change (Figure 1) aligns with the World Health Organization’s Behavioural and Social Drivers of Vaccination framework^25^ by targeting what people think and feel, social processes, and motivation or intention to vaccinate. The program trained “Vaccine Champions”, including health workers and diverse community leaders, to advocate for vaccines and address misinformation one-on-one and in groups. Vaccine Champions did not require health qualifications to participate and were selected by the Ministry of Health and UNICEF Vietnam if they had a trusted role in the community (e.g., village heads, leaders of women’s or youth groups) and were passionate about the health of their community. The training was co-designed with key stakeholders to provide comprehensive, tailored vaccine education and communication skills training appropriate for varied health literacy levels. The vaccines of focus in the program were identified during the initial co-design phase of the program and training included classroom-style education, interaction, role play, demonstrations, and activities, with modules on adult COVID-19 and routine childhood vaccines. Modules included information on vaccine-preventable diseases and vaccine development, safety, and effectiveness; vaccine communication techniques tailored across the vaccine acceptance spectrum; strategies for responding to misinformation and speaking one-on-one or in groups practised through role plays; instructions for organising an education session; and evaluation data collection.

**Figure 1:**
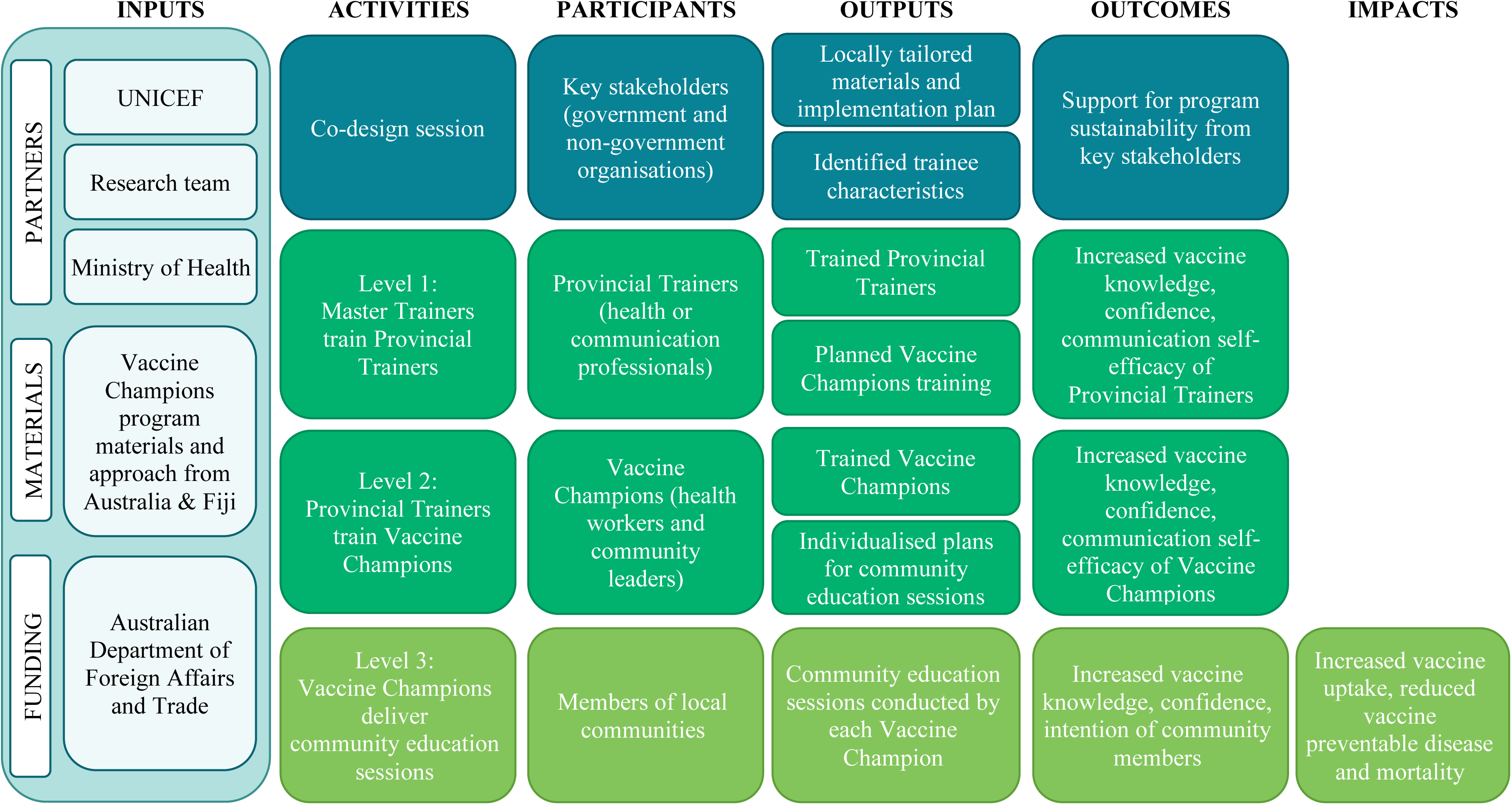
Theory of Change.

We utilised a Train-the-Trainer model with three levels for greater reach across Vietnam, which has a population of 100 million (Figure 2). Vietnamese experts in vaccination or communication provided input into training materials and were trained in English by the Australian academic team (JK, IO, MD) to be Master Trainers. The remainder of the program was conducted in Vietnamese with English simultaneous translation so researchers could provide continuous feedback and refinement. In Level 1, Master Trainers trained Provincial Trainers (healthcare workers and communication experts). In Level 2, Provincial Trainers trained health workers and community leaders to be Vaccine Champions. In Level 3, Vaccine Champions delivered education sessions to their communes. Those who attended sessions are termed “community attendees”, and those who responded to the survey are a subset of these, called “community respondents,” as not all attendees completed post-training surveys. Provincial Trainers and Vaccine Champions were provided travel, accommodation and per diems to attend training. Community attendees were given a token of appreciation such as food, toiletries or stationery. UNICEF provided logistical support for downstream trainings, education sessions and collection of evaluation data.

**Figure 2:**
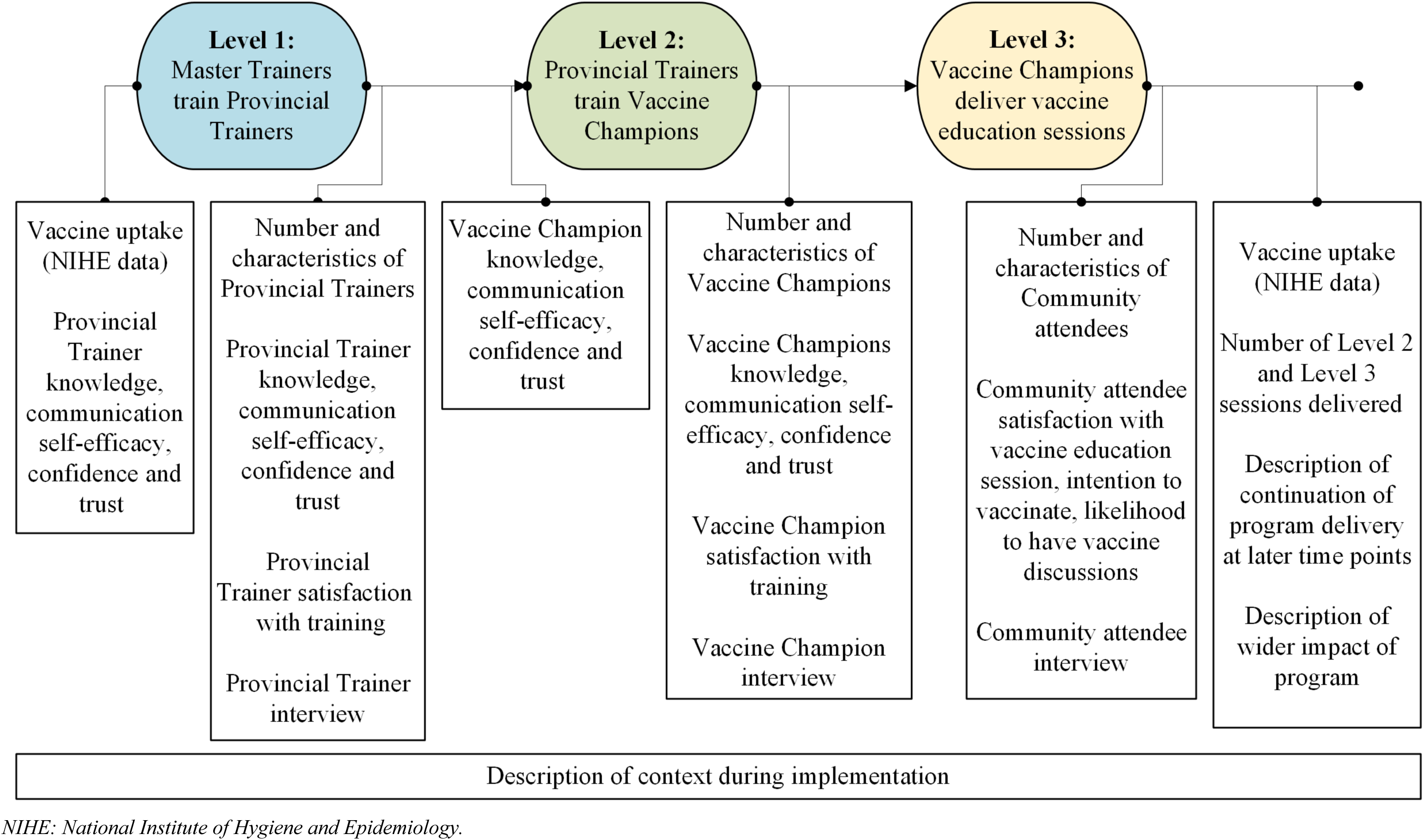
Training Levels and Data Collection Points.

### Pilot

The program was co-designed with Vietnamese government and stakeholders between August and November 2022 and piloted from 8-11 November 2022. Master Trainers conducted two Level 1 trainings in Gia Lai and Điện Biên, training 95 Provincial Trainers (55% female). Four Provincial Trainers from Điện Biên trained 32 Vaccine Champions (25% female) in a Level 2 training. Finally, four Champions conducted two Level 3 education sessions in Điện Biên, with 20 participants (25% female). All activities were observed and evaluated (Supplementary Tables 1-3). Following evaluation of the pilot, changes were made including increased training duration from one to one-and-a-half days; improvement and simplification of Vietnamese translations; inclusion of a guide for Vaccine Champions outlining key facts to share and reminding them to ask open questions in Level 3 sessions; simplification of data collection tools; additional content and activities about addressing misinformation; increased vaccine communication practice and feedback; reduced theory and more local examples and use of storytelling.

### Participants

All program participants who could consent and communicate in Vietnamese were eligible. The research was explained at the start of their first activity. Community attendees provided implied consent through optional, anonymous survey completion. All other participants provided written consent and data were collected during training or education sessions. The study team obtained consent from Provincial Trainers; Provincial Trainers obtained consent from Vaccine Champions; and Vaccine Champions explained implied consent if community attendees completed the survey.

### Data collection

We defined indicators to assess implementation outcomes according to RE-AIM categories (Table 1). We used interviews, surveys, and field notes, integrating qualitative, quantitative and observational data, to triangulate findings.

### Researcher field notes

Field notes were collected by researchers using a standard template, capturing data on reach, adoption and implementation. Adoption and maintenance data were provided by senior members of UNICEF Vietnam and Provincial Health Offices, who gave regular descriptive reports on downstream activities. We observed four Provincial Trainers trained in Sóc Trăng conduct a one-and-a-half-day Level 2 training and subsequently observed eight trained Vaccine Champions conduct two Level 3 sessions in their communes.

### Vaccine coverage reports

Provincial-level vaccine coverage was recorded in the three study provinces, Điện Biên, Sóc Trăng and Gia Lai, immediately pre- and 6-months post-intervention commencement. Measuring individual uptake was not feasible within resource constraints and was beyond the scope and funding for this program. As such, intention to vaccinate was a primary outcome, with provincial-level changes in coverage reported as a secondary outcome, noting diverse influences on uptake. The National Institute of Hygiene and Epidemiology provided COVID-19 booster dose vaccine data for individuals 18 years and over, compared to the province’s population. Data for ‘fully vaccinated’ children at 12 months according to the routine childhood schedule included children with: Bacille Calmette-Guérin (BCG) vaccine, 3 doses of diphtheria, tetanus, pertussis, hepatitis B, Haemophilus influenzae type B, at least three doses of polio (inactivated or oral) with at least one inactivated polio vaccine, and one measles. Percentages over 100% reflect a mobile population, causing the number of doses given to exceed the population number in some cases. Implementation provinces are graphically compared to control provinces in Supplementary Figure 1. Control provinces were selected if they were close to implementation provinces with similar geography.

### Surveys

Provincial Trainers and Vaccine Champions completed surveys immediately before and after training. The survey included items from validated tools and items generated by the research team to assess specific aspects of the program. Knowledge about vaccines and communication techniques were assessed by three questions developed by the research team, customised to the content of the training. Vaccine communication self-efficacy was assessed with four questions adapted from a validated communication self-efficacy scale for healthcare providers^26^ and trust and confidence in COVID-19 vaccines and the health system were assessed with two questions adapted from the validated World Health Organisation Behavioural and Social Drivers of Immunisation survey.^25^ Responses ranged from “not at all [confident]” to “very [confident]”. Data on demographics, COVID-19 vaccine uptake and intention to vaccinate were collected in the first survey, and training satisfaction in the second. Two questions were used to assess satisfaction, developed by our team, using a 3-point scale: not satisfied, satisfied, very satisfied.

Community attendees completed one survey with six questions immediately after an education session to assess change in vaccine confidence, communication and intention to receive COVID-19 vaccines for themselves or their children. Developed by the research team, questions had a 3-point scale of less, about the same, or more [confident or likely] than before attendance. We measured session satisfaction in one question using a 3-point response scale of unsatisfied, neutral, or satisfied and collected demographic data. Vaccine Champions reported how many community sessions they conducted and endeavoured to administer the survey to attendees after an education session. All items were reviewed by Vietnamese partners for face and content validity.

### Interviews

Group interviews were held after each observed session, exploring reach, effectiveness, implementation and maintenance. Participants were selected if they conducted training or were involved in training the next level. Six Provincial Trainers participated following the Level 1 training, four Provincial Trainers and eight Vaccine Champions participated following the Level 2 training, and four Provincial Trainers and seven Vaccine Champions participated following the Level 3 education session. Interviews followed semi-structured interview guides facilitated in person in English with a Vietnamese language translator, with audio recorded and transcribed.

### Data analysis

Survey data collected were categorical, with frequencies and percentages presented. Demographics and program satisfaction are summarised. Survey data from Provincial Trainers and Vaccine Champions were included in the effectiveness analysis where a response was given for both pre- and post-training surveys. Knowledge about vaccines and communication techniques are reported as percentages, with overall knowledge reported as the percentage of respondents who correctly answered all three knowledge questions. A sensitivity analysis was performed to explore the effect of assuming participants who did not answer a knowledge question did not know the answer, i.e. missing responses were treated as incorrect. Confidence items are reported as the percentage of respondents selecting “very confident”. Likewise, ‘trust in health system’ and ‘importance of a COVID-19 vaccine’ items are reported as the percentage of respondents selecting “very much”. McNemar’s test was used to compare pre- and post-training responses for binary variables. All analyses were unadjusted. A subgroup analysis for Vaccine Champions was performed comparing effectiveness by health background. For each subgroup, McNemar’s test was used to calculate the paired difference in post- minus pre-responses. Estimates are presented graphically. Analysis was performed using Stata version 18.0 (StataCorp, College Station, TX, USA, 2023).^27^

Qualitative data were collected to supplement the quantitative data in an explanatory sequential design.^28^ Interviews and field notes were analysed using directed qualitative content analysis.^29^ One author (IO) initially deductively coded data into the overarching RE-AIM categories of reach, effectiveness, adoption, implementation, and/or maintenance, as appropriate. Within each category, data were then grouped and summarised. Quantitative and qualitative results are integrated at the interpretation and reporting level through a narrative approach.^30^

### Ethical considerations

Approval was obtained from the Royal Children’s Hospital Human Research Ethics Committee (HREC:84863), and the Ethical Review Board for Biomedical Research at Hanoi University of Public Health (022-432/DD-YTCC).

### Public involvement

Immunisation stakeholders participated in the co-design workshop to inform intervention design. Health workers and community leaders were involved in intervention delivery as trainees and community members received the intervention.

## Results

The program was conducted from April 2023 to April 2024 in three low-coverage provinces.

### Reach

The program reached 56 Provincial Trainers (100% completed one survey and 82% (n=46) completed pre- and post-surveys) who trained 286 Vaccine Champions (73% (n=209) completed one survey and 71% (n=202) pre- and post-surveys). Table 2 describes characteristics of Provincial Trainers and Vaccine Champions.

**Table 2:** Baseline characteristics of Provincial Trainers and Vaccine Champions.

| Characteristic | Provincial Trainers (N=56) n (%) | Vaccine Champions (N=209) n (%) |
| --- | --- | --- |
| Education Level | N=55 | N=205 |
| High School or below | 1 (2) | 141 (69) |
| College, University or Vocational Education and Training | 43 (78) | 56 (27) |
| Post-Graduate (Masters or Doctor of Philosophy) | 11 (20) | 0 (0) |
| Other | 0 (0) | 8 (4) |
| Professional Role | N=56 | N=202 |
| Health<sup>Y</sup> | 37 (66) | 139 (69) |
| Communication | 14 (25) | N/A |
| Other<sup>f</sup> | 5 (9) | 63 (31) |
| Gender | N=55 | N=202 |
| Male | 29 (53) | 108 (54) |
| Female | 26 (47) | 93 (46) |
| Other or prefer not to say | 0 (0) | 1 (1) |
| Language | N=55 | N=199 |
| Vietnamese | 54 (98) | 169 (85) |
| Other<sup>#</sup> | 1 (2) | 30 (15) |
| Region | N=56 | N=204 |
| Northwest | 2 (4) | 71 (35) |
| Northeast | 1 (2) | 0 (0) |
| Red River Delta | 0 (0) | 0 (0) |
| North Central Coast | 1 (2) | 0 (0) |
| South Central Coast | 4 (7) | 0 (0) |
| Central Highlands | 8 (14) | 90 (44) |
| Southeast | 7 (13) | 4 (2) |
| Mekong River Delta | 33 (59) | 39 (19) |
| Number of COVID-19 vaccine doses received | N=56 | N=206 |
| 0 | 0 (0) | 2 (1) |
| 1 | 1 (2) | 0 (0) |
| 2 | 1 (2) | 2 (1) |
| 3 | 6 (11) | 35 (17) |
| More than 3 | 48 (86) | 167 (81) |
| Intention to receive another COVID-19 vaccine dose, if recommended | N=56 | N=204 |
| Yes | 47 (84) | 195 (96) |
| No | 4 (7) | 6 (3) |
| Not Sure | 5 (9) | 3 (1) |

<sup>#</sup>For Vaccine Champions, other languages spoken include Hmong or Mong, Khmer, Khmu (K'mu), Other, Xo Dang. <sup>Y</sup>For Provincial Trainers, health workers included professions such as doctors, nurses, pharmacists and village health workers. For Vaccine Champions, health workers included professions such as village health workers, health workers, and vaccination program staff. <sup>f</sup>For Vaccine Champions, 'other' included teachers, youth workers, police, community-based organisations (e.g. non-government organisations), government officials, Public Health students, tradespersons and faith leaders. N/A – not applicable. We asked the Vaccine Champions if they had a background in health or not, we did not give them a response option for 'communication'.

Vaccine Champions included health workers, village health workers, vaccination staff, teachers, youth workers, police, community-based organisations, government officials, public health students, tradespeople and faith leaders, with representation from all target groups achieved. Champions reached 4027 community attendees through education sessions in the study provinces and surrounding areas, with 28% (1134/4027) completing a post-session survey. Of these community respondents, 59% (666/1128) were female, and 93% (1020/1101) were from Điện Biên. Provincial Trainers and Vaccine Champions said the program was able to reach people from ethnic minorities and people living with disability by engaging Champions who were considered *“community volunteers”* who could go *“door-to-door as they know everybody in their village.”*

### Effectiveness

#### Vaccine knowledge

All Provincial Trainers and 94% of Vaccine Champions were correctly able to identify the vaccine not on the routine childhood schedule pre-training (see Table 3). Following training, there was no change in Provincial Trainer or Vaccine Champion knowledge regarding identification of correct statement about vaccines and correct communication technique to use, with 66% of Provincial Trainers and 31% of Vaccine Champions correctly identifying the vaccine communication technique post-training. Sensitivity analysis reached the same result (Supplementary Table 4 and Supplementary Figure 2). However, Provincial Trainers reflected that they learned new communication skills, and more detailed information about vaccines. Champions felt the training changed their approach to vaccine communication, with one saying:

> *“If we have a concern…from the community usually the habit is to address…questions right away, but in this course, it was designed to express our empathy with the questioner first and, you know, provide some further information before we address the issue… it will help us a lot in our future communication work.”*

**Table 3:**
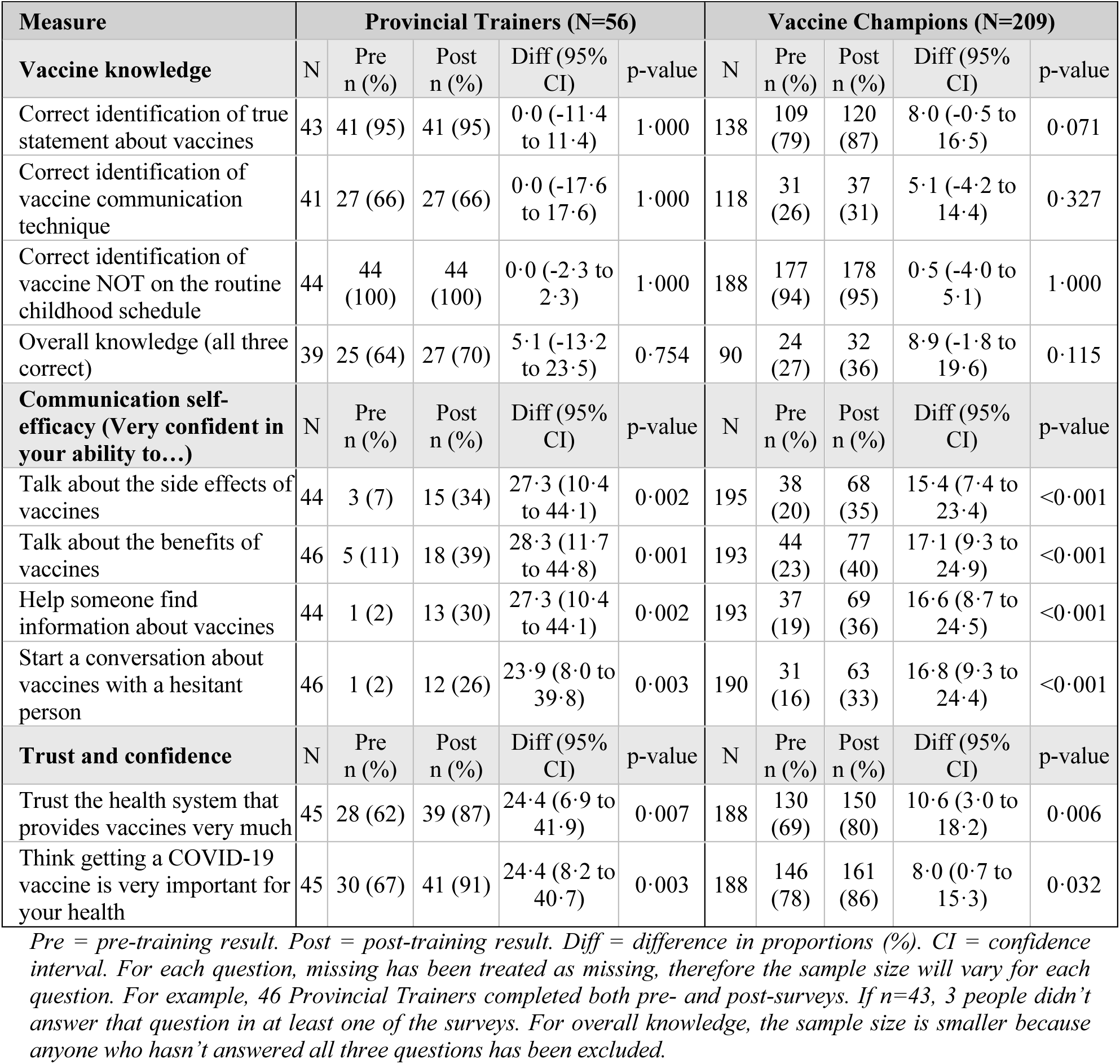
Changes in effectiveness outcomes for Provincial Trainers and Vaccine Champions.

| Measure | Provincial Trainers (N=56) |  |  |  |  | Vaccine Champions (N=209) |  |  |  |  |
| --- | --- | --- | --- | --- | --- | --- | --- | --- | --- | --- |
| Vaccine knowledge | N | Pre n (%) | Post n (%) | Diff (95% CI) | p-value | N | Pre n (%) | Post n (%) | Diff (95% CI) | p-value |
| Correct identification of true statement about vaccines | 43 | 41 (95) | 41 (95) | 0.0 (-11.4 to 11.4) | 1.000 | 138 | 109 (79) | 120 (87) | 8.0 (-0.5 to 16.5) | 0.071 |
| Correct identification of vaccine communication technique | 41 | 27 (66) | 27 (66) | 0.0 (-17.6 to 17.6) | 1.000 | 118 | 31 (26) | 37 (31) | 5.1 (-4.2 to 14.4) | 0.327 |
| Correct identification of vaccine NOT on the routine childhood schedule | 44 | 44 (100) | 44 (100) | 0.0 (-2.3 to 2.3) | 1.000 | 188 | 177 (94) | 178 (95) | 0.5 (-4.0 to 5.1) | 1.000 |
| Overall knowledge (all three correct) | 39 | 25 (64) | 27 (70) | 5.1 (-13.2 to 23.5) | 0.754 | 90 | 24 (27) | 32 (36) | 8.9 (-1.8 to 19.6) | 0.115 |
| Communication self-efficacy (Very confident in your ability to...) | N | Pre n (%) | Post n (%) | Diff (95% CI) | p-value | N | Pre n (%) | Post n (%) | Diff (95% CI) | p-value |
| Talk about the side effects of vaccines | 44 | 3 (7) | 15 (34) | 27.3 (10.4 to 44.1) | 0.002 | 195 | 38 (20) | 68 (35) | 15.4 (7.4 to 23.4) | <0.001 |
| Talk about the benefits of vaccines | 46 | 5 (11) | 18 (39) | 28.3 (11.7 to 44.8) | 0.001 | 193 | 44 (23) | 77 (40) | 17.1 (9.3 to 24.9) | <0.001 |
| Help someone find information about vaccines | 44 | 1 (2) | 13 (30) | 27.3 (10.4 to 44.1) | 0.002 | 193 | 37 (19) | 69 (36) | 16.6 (8.7 to 24.5) | <0.001 |
| Start a conversation about vaccines with a hesitant person | 46 | 1 (2) | 12 (26) | 23.9 (8.0 to 39.8) | 0.003 | 190 | 31 (16) | 63 (33) | 16.8 (9.3 to 24.4) | <0.001 |
| Trust and confidence | N | Pre n (%) | Post n (%) | Diff (95% CI) | p-value | N | Pre n (%) | Post n (%) | Diff (95% CI) | p-value |
| Trust the health system that provides vaccines very much | 45 | 28 (62) | 39 (87) | 24.4 (6.9 to 41.9) | 0.007 | 188 | 130 (69) | 150 (80) | 10.6 (3.0 to 18.2) | 0.006 |
| Think getting a COVID-19 vaccine is very important for your health | 45 | 30 (67) | 41 (91) | 24.4 (8.2 to 40.7) | 0.003 | 188 | 146 (78) | 161 (86) | 8.0 (0.7 to 15.3) | 0.032 |

*Pre = pre-training result. Post = post-training result. Diff = difference in proportions (%). CI = confidence interval. For each question, missing has been treated as missing, therefore the sample size will vary for each question. For example, 46 Provincial Trainers completed both pre- and post-surveys. If n=43, 3 people didn't answer that question in at least one of the surveys. For overall knowledge, the sample size is smaller because anyone who hasn't answered all three questions has been excluded.*

After the Level 3 education session, Provincial Trainers reflected on Vaccine Champions’ performance, noting *“communication skills are very good”*, but vaccine knowledge could be improved. Provincial Trainers said for future Level 2 training, they will *“focus more on knowledge”* that aligns with changing community concerns about vaccines. When interviewed after the Level 2 training, Vaccine Champions commented they would be *“very confident talking about safety with the community”*, would try to answer concerns coming from their communities, and knew to refer to a Provincial Trainer if they didn’t know the answer. Champions also commented while *“everything was understandable and we absorbed all information provided by the training”,* they desired longer trainings to include more information on childhood vaccines.

#### Communication self-efficacy, trust and confidence

Communication self-efficacy increased significantly across all measures among Provincial Trainers and Vaccine Champions (see Table 3). The percentage of respondents in both groups who were very confident discussing vaccine benefits and side effects, helping someone access information about vaccines and starting a conversation with a hesitant person increased post-training. The increase was greater amongst Provincial Trainers, ranging from 24% to 28%, compared to Vaccine Champions. The percentage of respondents in both groups who trusted the health system and saw the COVID-19 vaccine as important for their health also increased.

Changes in effectiveness outcomes for Vaccine Champions with a health and non-health background are seen in Figure 3, with improvement in most topics post training.

**Figure 3:**
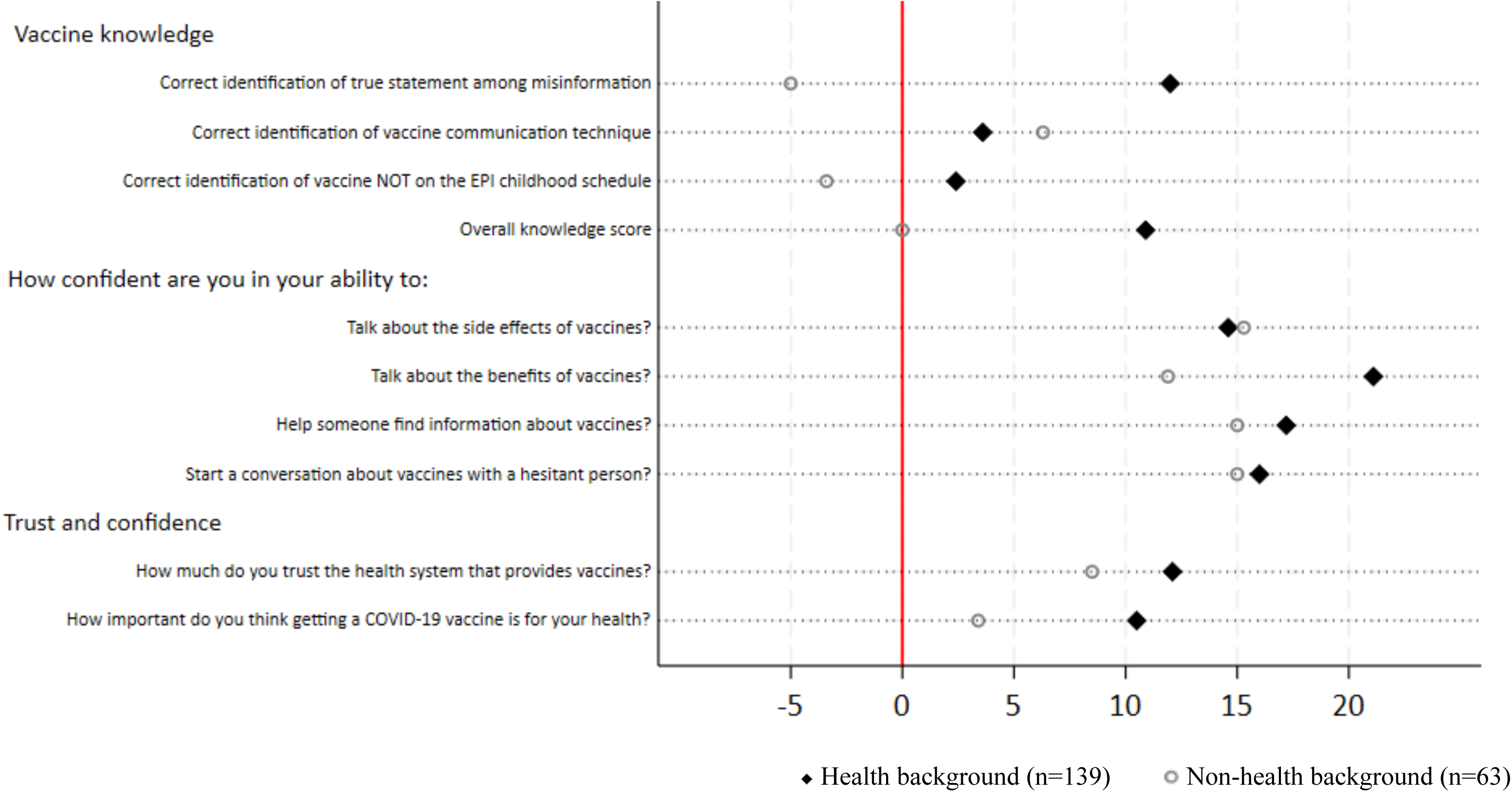
Percentage difference in effectiveness outcomes (post-training compared to pre-training) for Vaccine Champions with a health and non-health background.

#### Intention to vaccinate

Among community respondents (28% of attendees), the majority were more likely to get a COVID-19 vaccine for themselves (964/1121, 86%) and their child (966/1073, 90%), than before the session. Most were also more confident that routine vaccines were important for children (994/1121, 89%), and that COVID-19 vaccines were important for themselves (1042/1123, 93%), and children (998/1115, 90%) post the session.

#### Satisfaction

All Provincial Trainers and Vaccine Champions reported they were satisfied or very satisfied with the training and materials provided (Supplementary Figure 3). Most community respondents were also satisfied with the session (1020/1124, 91%).

#### Vaccine coverage

Adult COVID-19 booster and childhood vaccine coverage at provincial level increased in each implementation province between April and October 2023. COVID-19 booster coverage increased from 27% to 94% in Điện Biên, 6% to 101% in Sóc Trăng, and 34% to 72% in Gia Lai and childhood vaccine coverage increased from 21% to 45% in Điện Biên, 15% to 62% in Sóc Trăng, and 16% to 34% in Gia Lai. Coverage also increased in adjacent control regions, although less marked (Supplementary Figure 1).

### Adoption

#### Number of trainings and sessions

Between 1 April 2023 and 1 April 2024, Provincial Trainers conducted ten Vaccine Champions trainings in Level 2 and Vaccine Champions conducted 190 education sessions in Level 3.

#### Likelihood of initiating vaccine discussions with social networks

Most community respondents reported they were more likely to talk about vaccines with other people after an education session (87%, 982/1123), with 10% reporting no change (115/1123), and 2% being less likely (26/1123).

### Implementation

#### Program adaptations

Provincial Trainers suggested when delivering a future Level 2 training, they would expand content about childhood vaccines, remove a vaccine communication activity about designing a vaccine message, and deliver the training in pairs using *“one person with good knowledge, and one person with good communication skills”*. Provincial Trainers reported they didn’t have enough time to cover all the content during training, but did make time to answer questions, which they found *“motivated participants to join in”*.

Vaccine Champions said they would *“localise the language”* in materials to suit the ethnic minority groups they served. They planned to invite attendees to their education sessions using Zalo (a mobile communication application), the local loudspeaker, and calling individuals. After sessions Champions reflected *“all community concerns were addressed fully with confidence”*, but they would have liked additional materials and information to support their session, with more detail, frequently asked questions, and culturally appropriate images. Vaccine Champions were able to tailor sessions to suit their audience, such as focusing only on COVID-19 boosters when parents did not attend.

#### Local context during implementation

Throughout Vietnam, there were childhood vaccine supply issues immediately prior to program commencement, and extreme flooding from August 2023.^31–34^ In Gia Lai and Điện Biên, flooding led to dangerous landslides.^31, 34^ In Gia Lai and Điện Biên, rabies cases were recorded in 2023 with 14 and six deaths, respectively, resulting in nearly half a million people receiving a rabies vaccine at this time.^35^ Điện Biên recorded six diphtheria cases in 2023, including one death, which was followed by a push for vaccination against diphtheria.^36^In Gia Lai in June, there was a gun attack which resulted in a halt in our program’s downstream activities.^37^

### Maintenance

#### Continuation of program delivery

Since the end of the study period in April 2024, the program continues with government support. Provincial Trainers have run an additional 14 Vaccine Champions trainings, reaching 427 participants across Gia Lai, Điện Biên and Sóc Trăng between May and December 2024. Additionally, Vaccine Champions ran 210 community education sessions in Gia Lai reaching 4,312 attendees; 90 sessions in Điện Biên reaching 2,340 attendees; and 48 sessions in Sóc Trăng reaching 1440 attendees.

#### Program influence and sustainability

Champions reflected that the communication skills they learned were *“flexible to apply to other topics”* such as dengue prevention. Champions from women’s groups mentioned it would be easy to *“integrate vaccination [education] into their routine monthly meetings”*.

Champions have since been engaged by UNICEF and the Ministry of Health to communicate about other health issues, including nutrition, and maternal and child health. Training materials have been utilised by UNICEF and the Ministry of Health to train Vietnamese journalists to communicate about vaccines. At the end of the study period, in collaboration with the National Institute of Hygiene and Epidemiology, a vaccine information and communication handbook was developed to support health workers and trainees to communicate about vaccines.^35^ This has been used by immunisation officers, responsible for immunisation activities, across all provinces in Vietnam.

Both Provincial Trainers and Vaccine Champions reflected that while most hesitancy was related to COVID-19 vaccines, there was some hesitancy surrounding childhood vaccines, particularly following adverse events. They commented that non-health Champions should provide support to health workers to communicate more broadly about vaccine safety, *“…so we need to have these kinds of [non-health] Champions, they need to be provided with the skill and they can help the health workforce in case of responding to the hesitancy with the EPI [Expanded Program on Immunisation]-related event”*.

## Discussion

Our Train-the-Trainer Vaccine Champions program, conducted in three provinces in Vietnam between April 2022 and April 2023, engaged health workers and community leaders to promote COVID-19 and childhood vaccines during the pandemic. Codesigned with the Ministry of Health, National Institute of Hygiene and Epidemiology, and UNICEF, the program was piloted and tailored to community needs. Communication self-efficacy, trust in vaccines and the health system and intention to vaccinate increased, although change in vaccine knowledge was minimal. All participant groups reported high satisfaction with sessions and materials. Champions reached over 4000 community attendees across Vietnam during the program period and reached an additional 8000 attendees in 2023 and 2024. The program has ongoing support from the Ministry of Health and Vaccine Champions have been utilised to advocate for other health issues in communities in Vietnam.

Effective, targeted communication about vaccines is needed globally to address declining trust and confidence in routine vaccines.^38^ Health worker training to improve confidence in communicating about vaccine side effects, answering questions or having conversations with hesitant people is recommended in many settings.^39–44^ However, evidence for interventions aimed at healthcare workers is inconsistent, with some studies indicating effectiveness^18, 45–47^ while others find no impact.^26, 48^ Evidence of vaccine communication training interventions among non-health workers is limited. Our program specifically targets community members as well as health workers with comprehensive capacity building in both vaccine knowledge and communication skills, tailored to the communicator’s previous level of health training. While confidence increased in our study, objective knowledge measures did not increase in either group after our training, potentially due in part to the relatively high knowledge levels pre-training, particularly among Provincial Trainers. However, we used these findings to subsequently develop a vaccine education and communication handbook with the National Institute of Hygiene and Epidemiology, which now supports Champions and has been reported as a valuable resource.^49^

Training both health and non-health Champions has many advantages. Champions with a health background are highly trusted and generally improve more significantly with training due to higher baseline education levels and familiarity with training approaches. However, vaccine messages are strengthened when shared by community leaders, utilising an intersectoral approach.^50–52^ Community Champions can relieve healthcare worker burden, with effectiveness demonstrated for other health promotion activities in Vietnam,^51^as well as globally.^18, 19^ Community Champions are particularly valuable in accessing remote locations underserved by health staff^53^ and in reaching under-vaccinated people from diverse ethnic backgrounds, especially if they are from these communities and know the local traditions and languages needed to communicate effectively and build trust. Our program reached communities in northern, central and southern Vietnam and supported community Champions from distant communes to attend training sessions before returning to share what they had learnt with their local communities. In addition, our program had a strong gender lens, prioritising inclusion of women at all levels. To build trust in vaccines and minimise the spread of misinformation, conversations about vaccines need to reach fathers, mothers and extended family members to address gender disparities and reach the primary decision-maker in the household.^54, 55^

In our study, community attendees reported an increased likelihood to talk to their social networks about vaccines after education sessions, suggesting a broader ripple effect of the program beyond those receiving formal education sessions. When there are threats to community health, people are more motivated to talk to one another, seek trustworthy information and accept and act on vaccination recommendations from people they know and trust.^56, 57^ Empowering grass roots advocacy and shaping social norms through face-to-face and online interpersonal communication training to prevent infectious disease outbreaks has been effective before in Vietnam.^58^ Face-to-face education has also been shown to be effective in other preventative health programs in Vietnam, including disease control,^52^ drug use,^59^ food hygiene,^60^ eye health,^61^ and with the ageing population.^62^

Intention to vaccinate increased among community attendees but impact on individual vaccine uptake was not assessed due to resource constraints during the COVID-19 pandemic and the pragmatic nature of the study. Whilst increased confidence in vaccines and intention are strongly linked to uptake, ^63^ there is an intention-behaviour gap. However, intention to vaccinate is a valid assessment of the effectiveness of a face-to-face knowledge and communication skills intervention as uptake is often moderated by access and supply issues that are not targeted by this intervention. Additionally, improvements in trust in vaccines and healthcare systems, vaccine knowledge, and communication skills are valuable real word outcomes.^64^ Although coverage increased in intervention provinces, we were unable to isolate the intervention’s impact on coverage at the provincial level due to other strategies to improve uptake being introduced concurrently.

Our study’s strengths include engagement with the Ministry of Health and UNICEF, a comprehensive multi-level training program in three provinces, and evaluation of implementation outcomes using the RE-AIM framework. However, there were several limitations. Many Champions did not complete all knowledge questions, possibly indicating that adjustments are needed to better evaluate knowledge. Only a quarter of attendees completed the community attendee survey, which was optional to reduce the burden on community Champions. This may have introduced response and social desirability bias. As outlined, individual coverage data collection was beyond the scope of the study and provincial level coverage was a secondary outcome. Individual level vaccine uptake needs to be assessed in future hybrid implementation-effectiveness trials. Collection of economic data is needed in the future to guide equitable adoption and scalability. Future studies should also incorporate access strategies, such as immunisation outreach, to address both acceptance and behaviour, reflected by vaccine uptake.

## Conclusion

Our comprehensive, multi-level Vaccine Champions program trained health and community leaders to promote routine childhood and COVID-19 vaccination, resulting in increased trust in vaccines and the health system, communication self-efficacy, and community intention to vaccinate in Vietnam. The program has potential to reduce healthcare workforce burden and has ongoing government support, with Champions being utilised to promote vaccines and other health priorities including nutrition and maternal and child health in Vietnam. Hybrid implementation-effectiveness trials are now needed to determine impact on vaccine coverage and cost-effectiveness, alongside implementation outcomes, to guide scalability across Vietnam and adaption in other countries.

## Data Availability

De-identified individual participant data that underlie the results reported in this article, as well as the study protocol and data collection instruments, will be made available from the point of, and up to three years after the acceptance for publication of the main findings. Data will be shared with researchers who provide a methodologically sound proposal, for analyses that achieve the aims in the approved proposal. Proposals should be directed to the corresponding author. To gain access, data requesters will need to sign a data access agreement.

## Abbreviations

BCG: Bacille Calmette-Guérin
COVID-19: Coronavirus Disease 2019
EPI: Expanded Program on Immunisation
HREC: Human Research Ethics Committee
N/A: Not Applicable
NIHE: National Institute of Hygiene and Epidemiology
RE-AIM: Reach, Effectiveness, Adoption, Implementation, and Maintenance
SDG3: Sustainable Development Goal Three
STROBE: Strengthening the Reporting of Observational Studies in Epidemiology
UNICEF: United Nations Children’s Fund

## Declarations

## Acknowledgements

Thank you to the Vietnamese Ministry of Health and the National Institute of Hygiene and Epidemiology, for their support of the program. Thank you to the participants who gave up their time to contribute to the program and research. Thank you to Jaysha Tippins for supporting data entry.

## Ethics approval and consent to participate

Approval was obtained from the Royal Children’s Hospital Human Research Ethics Committee (HREC:84863), and the Ethical Review Board for Biomedical Research at Hanoi University of Public Health (022-432/DD-YTCC). Community attendees provided implied consent through survey completion. All other participants provided written consent.

## Authors’ contributions

Margie Danchin, Jessica Kaufman, Gregory Fox, Thu-Anh Nguyen, Maharajan Muthu, Chu Huu Trang, Shiva Shrestha, Holly Seale and Isabella Overmars were involved in study conception. Margie Danchin, Jessica Kaufman, Gregory Fox, Thu-Anh Nguyen, Maharajan Muthu, Chu Huu Trang, Shiva Shrestha and Holly Seale were involved in funding acquisition. All authors were involved in design of the program and/or evaluation. Maharajan Muthu, Chu Huu Trang, Thu-Anh Nguyen, Luong Tran, Thi Mai Nguyen, Vu Nguyen Ngoc Anh, Jessica Kaufman, Isabella Overmars and Margie Danchin made significant contributions to program implementation. Gregory Fox, Thu-Anh Nguyen, Maharajan Muthu, Shiva Shrestha and Margie Danchin contributed to the project’s administration. Isabella Overmars, Jessica Kaufman, Chu Huu Trang, Thu-Anh Nguyen, Luong Tran, Thi Mai Nguyen, and Vu Nguyen Ngoc Anh contributed to data collection. Isabella Overmars, Suzanna Vidmar, Jessica Kaufman and Margie Danchin contributed to data analysis. Isabella Overmars wrote the original draft. Jessica Kaufman and Margie Danchin primarily reviewed and edited the original draft. All authors were involved in interpretation of data and critical revision of the manuscript.

## Availability of data and materials

De-identified individual participant data that underlie the results reported in this article, as well as the study protocol and data collection instruments, will be made available from the point of, and up to three years after the acceptance for publication of the main findings. Data will be shared with researchers who provide a methodologically sound proposal, for analyses that achieve the aims in the approved proposal. Proposals should be directed to. To gain access, data requesters will need to sign a data access agreement.

