## Supplementary Data for "Pragmatic RE-AIM evaluation of the Train-the-Trainer Vaccine Champions Program in Vietnam"

### Pilot Data - November 2022

95 Provincial Trainers participated in the survey. 89 completed both pre- and post-training surveys. 32 Vaccine Champions participated in the survey, and all completed both pre- and post-surveys. We did not collect data from the community attendees who received an education session.

#### Supplementary Table 1: Pilot: Characteristics of Provincial Trainers and Vaccine Champions

| Characteristic | Provincial Trainers (N=95) n (%) | Vaccine Champions (N=32) n (%) |
|---|---|---|
| Education Level | N=95 | N=32 |
| High School or below | 0 (0) | 14 (44) |
| College, University or Vocational Education and Training | 67 (71) | 17 (53) |
| Post-Graduate (Masters or Doctor of Philosophy) | 27 (28) | 1 (3) |
| Other | 1 (1) | 0 (0) |
| Professional Role | N=95 | N=32 |
| Health[Y] | 66 (70) | 13 (41) |
| Communication | 28 (30) | N/A |
| Other[f] | 1 (1) | 19 (59) |
| Gender | N=94 | N=32 |
| Male | 41 (44) | 24 (75) |
| Female | 52 (55) | 8 (25) |
| Other or prefer not to say | 1 (1) | 0 (0) |
| Language | N=94 | N=32 |
| Vietnamese | 94 (100) | 14 (44) |
| Other | 0 (0) | 18 (56) |
| Region | N=94 | N=31 |
| Northwest | 22 (23) | 31 (100) |
| Northeast | 21 (22) | 0 (0%) |
| Red River Delta | 0 (0) | 0 (0%) |
| North Central Coast | 20 (21) | 0 (0%) |
| South Central Coast | 12 (13) | 0 (0%) |
| Central Highlands | 19 (20) | 0 (0%) |
| Southeast | 0 (0) | 0 (0%) |
| Mekong River Delta | 0 (0) | 0 (0%) |

[Y] *For Provincial Trainers, health workers included professions such as doctors, nurses and pharmacist. For Vaccine Champions, health workers included professions such as village health workers, health workers, and vaccination program staff. N/A – not applicable. We asked the Vaccine Champions if they had a background in health or not, we did not give them a response option for 'communication'.*

### Supplementary Table 2: Pilot: Effectiveness outcomes for Provincial Trainers and Vaccine Champions

| Measure | Provincial Trainers (N=89) | | | | | Vaccine Champions (N=32) | | | | |
|---|---|---|---|---|---|---|---|---|---|---|
| | N | Pre | Post | Diff (95% CI) | p-value | N | Pre | Post | Diff (95% CI) | p-value |
| *Vaccine knowledge: correct identification of COVID-19 vaccine side effects, n(%)* | | | | | | | | | | |
| Side effect - Sore arm | 89 | 88 (99) | 88 (99) | 0·0 (-4·2 to 4·2) | 1·000 | 32 | 24 (75) | 29 (91) | 15·6 (-2·8 to 34·0) | 0·125 |
| Side effect - Headache | 89 | 60 (67) | 81 (91) | 23·6 (12·6 to 34·6) | <0·001 | 32 | 15 (47) | 25 (78) | 31·3 (7·9 to 54·6) | 0·013 |
| Side effect - Body ache | 89 | 61 (69) | 76 (85) | 16·9 (5·8 to 27·9) | 0·003 | 32 | 15 (47) | 17 (53) | 6·3 (-8·9 to 21·4) | 0·625 |
| Side effect - Mild fever | 89 | 85 (96) | 89 (100) | 4·5 (-0·9 to 9·9) | 0·125 | 32 | 31 (97) | 32 (100) | 3·1 (-6·0 to 12·3) | 1·000 |
| Side effect - Fatigue | 89 | 83 (93) | 86 (97) | 3·4 (-3·5 to 10·3) | 0·453 | 32 | 24 (75) | 29 (91) | 15·6 (-0·1 to 31·3) | 0·063 |
| *Vaccine knowledge: correct identification of diseases not on the routine vaccination schedule, n(%)* | | | | | | | | | | |
| Not on routine vaccination schedule - Malaria | 89 | 73 (82) | 77 (87) | 4·5 (-4·2 to 13·2) | 0·388 | 32 | 20 (63) | 18 (56) | -6·2 (-32·2 to 19·7) | 0·791 |
| Not on routine vaccination schedule - HIV | 89 | 79 (89) | 84 (94) | 5·6 (-2·0 to 13·2) | 0·180 | 32 | 16 (50) | 20 (63) | 12·5 (-5·0 to 30·0) | 0·219 |
| *Communication skill knowledge: correct identification of recommended communication techniques, n(%)* | | | | | | | | | | |
| Strategy - Talk about the risks of COVID-19 disease | 89 | 58 (65) | 63 (71) | 5·6 (-6·9 to 18·1) | 0·442 | 32 | 9 (28) | 14 (44) | 15·6 (-12·2 to 43·4) | 0·332 |
| Strategy - Recommend vaccination | 89 | 69 (78) | 82 (92) | 14·6 (3·9 to 25·4) | 0·007 | 32 | 9 (28) | 20 (63) | 34·4 (12·7 to 56·1) | 0·003 |
| Strategy - Share your experiences | 89 | 73 (82) | 88 (99) | 16·9 (7·4 to 26·4) | <0·001 | 32 | 19 (60) | 25 (78) | 18·8 (-4·6 to 42·1) | 0·146 |
| Strategy - Ask what concerns the person has | 89 | 78 (88) | 87 (98) | 10·1 (1·3 to 18·9) | 0·022 | 32 | 18 (56) | 25 (78) | 21·9 (-0·1 to 43·8) | 0·065 |
| Strategy - Continue the conversation | 89 | 61 (69) | 75 (84) | 15·7 (3·9 to 27·6) | 0·009 | 32 | 8 (25) | 12 (38) | 12·5 (-7·4 to 32·4) | 0·289 |
| *Trust and confidence in COVID-19 vaccines (very much), n(%)* | | | | | | | | | | |
| Trust the health system | 88 | 54 (61) | 69 (78) | 17·0 (6·9 to 27·2) | 0·001 | 32 | 16 (50) | 16 (50) | 0·0 (-18·1 to 18·1) | 1·000 |
| Trust COVID-19 vaccines | 89 | 48 (54) | 64 (72) | 18·0 (6·7 to 29·2) | 0·002 | 32 | 17 (53) | 16 (50) | -3·1 (-24·6 to 18·3) | 1·000 |
| Think getting a COVID-19 vaccine is important for your health | 86 | 53 (62) | 67 (78) | 16·3 (5·5 to 27·0) | 0·003 | 32 | 19 (59) | 15 (47) | -12·5 (-38·1 to 13·1) | 0·424 |
| Think getting a COVID-19 vaccine will protect others | 87 | 48 (55) | 61 (70) | 14·9 (2·5 to 27·4) | 0·019 | 31 | 14 (45) | 15 (48) | 3·2 (-18·9 to 25·4) | 1·000 |
| Think a COVID-19 vaccine is safe for you | 88 | 33 (38) | 56 (64) | 26·1 (15·3 to 37·0) | <0·001 | 31 | 12 (39) | 10 (32) | -6·5 (-25·0 to 12·1) | 0·688 |
| Think COVID-19 vaccines work well to prevent disease | 89 | 33 (37) | 56 (63) | 25·8 (14·1 to 37·5) | <0·001 | 32 | 12 (38) | 16 (50) | 12·5 (-14·7 to 39·7) | 0·454 |
| *Communication self-efficacy (0 cannot do at all – 10 highly certain I can do), mean (SD)* | | | | | | | | | | |
| Talk about the side effects of vaccines | 81 | 6·6 (1·9) | 8·4 (1·4) | 1·9 (1·5 to 2·3) | <0·001 | 26 | 4·9 (1·9) | 7·0 (1·8) | 2·1 (1·0 to 3·1) | <0·001 |
| Talk about the benefits of vaccines | 87 | 6·9 (2·1) | 8·5 (1·4) | 1·6 (1·3 to 2·0) | <0·001 | 31 | 6·5 (2·5) | 7·9 (1·8) | 1·4 (0·4 to 2·3) | 0·007 |
| Help someone find information about vaccines | 86 | 6·9 (2·0) | 8·5 (1·5) | 1·6 (1·2 to 1·9) | <0·001 | 30 | 6·2 (2·6) | 7·1 (2·0) | 1·0 (0·0 to 1·9) | 0·043 |
| Answer questions about vaccines | 87 | 6·6 (2·1) | 8·5 (1·4) | 1·9 (1·5 to 2·2) | <0·001 | 32 | 5·5 (2·4) | 7·4 (1·7) | 1·9 (1·2 to 2·7) | <0·001 |
| Start a conversation about vaccines with a hesitant person | 86 | 6·6 (2·2) | 8·6 (1·4) | 2·0 (1·6 to 2·4) | <0·001 | 32 | 6·1 (2·6) | 7·3 (2·0) | 1·2 (0·2 to 2·2) | 0·020 |
| Address misinformation about vaccines | 85 | 6·3 (2·2) | 8·4 (1·5) | 2·1 (1·7 to 2·6) | <0·001 | 32 | 5·6 (2·5) | 7·7 (1·9) | 2·1 (1·0 to 3·1) | <0·001 |

*^Total matched pre- and post-training responses for each question vary due to missing data. Categorical responses are presented as numbers and percentages. Knowledge is reported as the percentage of participants who correctly identified side effects, recommended communication techniques, and childhood vaccines. Vaccine trust and confidence is reported as the percentage of participants responding, "very much". Satisfaction is reported as the percentage of participants responding, "very satisfied". Communication self-efficacy was analysed as a continuous variable with mean scores and standard deviation (SD) presented. McNemar's test was used to compare pre- and post-training responses for binary variables while the paired t test was used to compare continuous variables. Data from pre- and post-training surveys that could not be matched to a single participant due to labelling or survey completion errors are not included in the analysis. Analysis was performed using Stata version 18.0. Routine vaccines are Expanded Program on Immunisation (EPI) vaccines. COVID-19: Coronavirus disease 2019. HIV: Human immunodeficiency virus.*

### Supplementary Table 3: Pilot: Intervention Satisfaction of Provincial Trainers and Vaccine Champions

| ***How satisfied were you with:*** | **Not satisfied** ***n (%)*** | **Satisfied** ***n (%)*** | **Very satisfied** ***n (%)*** |
|---|---|---|---|
| **Level 1, Provincial Trainers** | | | |
| The training session you attended (n=89) | 0 (0) | 20 (23) | 69 (78) |
| The materials provided to you for running your own session (n=89) | 0 (0) | 25 (28) | 64 (72) |
| **Level 2, Vaccine Champions** | | | |
| The training session you attended (n=32) | 0 (0) | 8 (25) | 24 (75) |
| The materials provided to you for running your own session (n=30) | 0 (0) | 13 (43) | 17 (57) |

### Vaccine Coverage in Implementation and Control Provinces

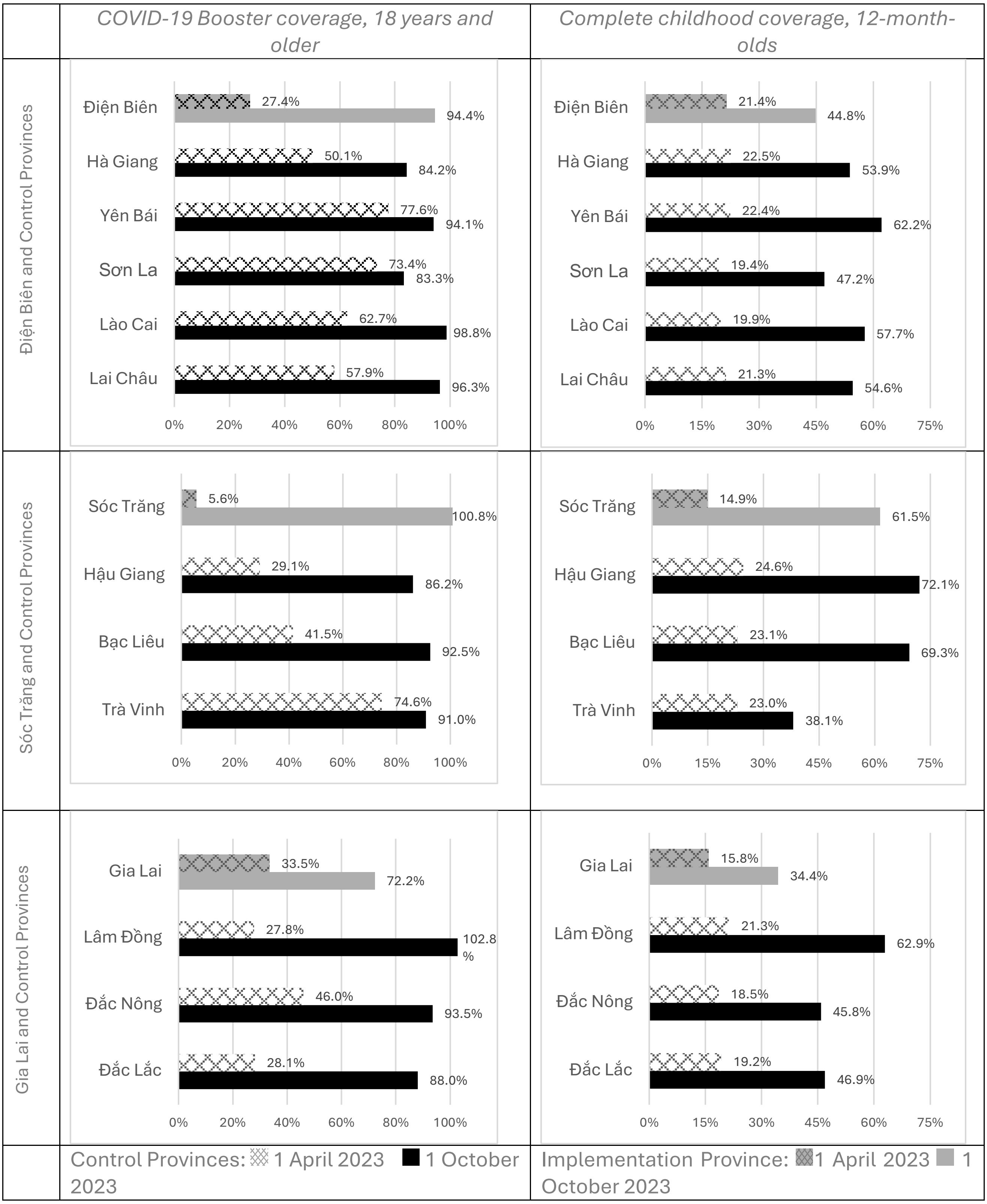


**Supplementary Figure 1: COVID-19 booster and routine childhood vaccine coverage in Điện Biên, Sóc Trăng, Gai Lai and control provinces.** *Where percentages are listed are over 100%, this is due to mobile populations in some provinces, which led to the number of actual doses given being higher than the total population estimate for a specific province, according to data from the National Institute of Hygiene and Epidemiology (NIHE).*

#### Sensitivity Analysis – Training Effectiveness on Knowledge

For this analysis, we treated missing knowledge data as incorrect.

**Supplementary Table 4: Sensitivity Analysis: Changes in effectiveness on knowledge outcomes for Provincial Trainers and Vaccine Champions**

| Measure | Provincial Trainers (N=56) | | | | | Vaccine Champions (N=209) | | | | |
|---|---|---|---|---|---|---|---|---|---|---|
| **Vaccine knowledge** | N | Pre n (%) | Post n (%) | Diff (95% CI) | p-value | N | Pre n (%) | Post n (%) | Diff (95% CI) | p-value |
| Correct identification of true statement about vaccines | 46 | 41 (89.1) | 43 (93.5) | 4.3 (-8.2 to 16.9) | 0.688 | 202 | 129 (63.9) | 142 (70.3) | 6.4 (-2.3 to 15.2) | 0.160 |
| Correct identification of vaccine communication technique | 46 | 28 (60.9) | 29 (63.0) | 2.2 (-15.3 to 19.7) | 1.000 | 202 | 36 (17.8) | 44 (21.8) | 4.0 (-2.5 to 10.4) | 0.256 |
| Correct identification of vaccine NOT on the routine childhood schedule | 46 | 44 (95.7) | 46 (100.0) | 4.3 (-3.7 to 12.4) | 0.500 | 202 | 178 (88.1) | 186 (92.1) | 4.0 (-1.3 to 9.2) | 0.152 |
| Overall knowledge (all three correct) | 46 | 26 (56.5) | 29 (63.0) | 6.5 (-10.9 to 23.9) | 0.581 | 202 | 28 (13.9) | 37 (18.3) | 4.5 (-1.2 to 10.1) | 0.136 |

*Pre = pre-training result. Post = post-training result. Diff = difference in proportions (%). CI = confidence interval. For each question, missing has been treated as incorrect.*

**Supplementary Figure 2: Sensitivity Analysis: Percentage difference in effectiveness outcomes for Vaccine Champions with a health and non-health background.**

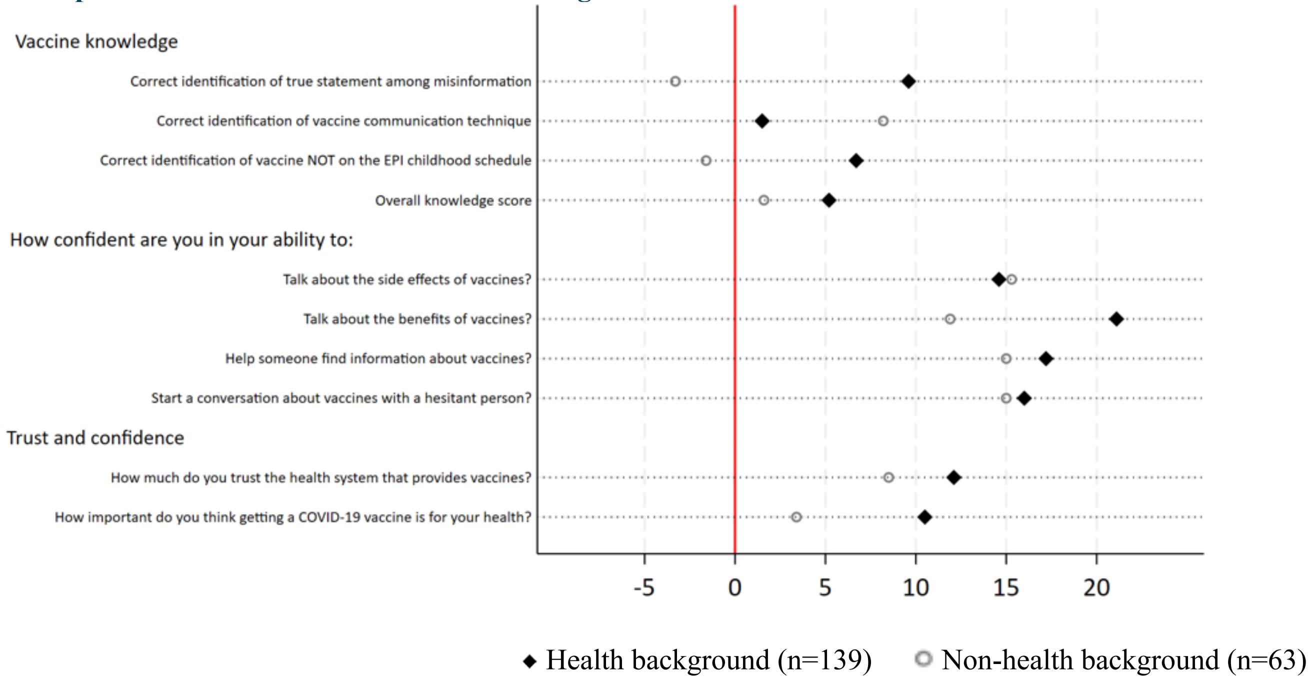


*For each knowledge question, missing has been treated as incorrect.*

### Satisfaction

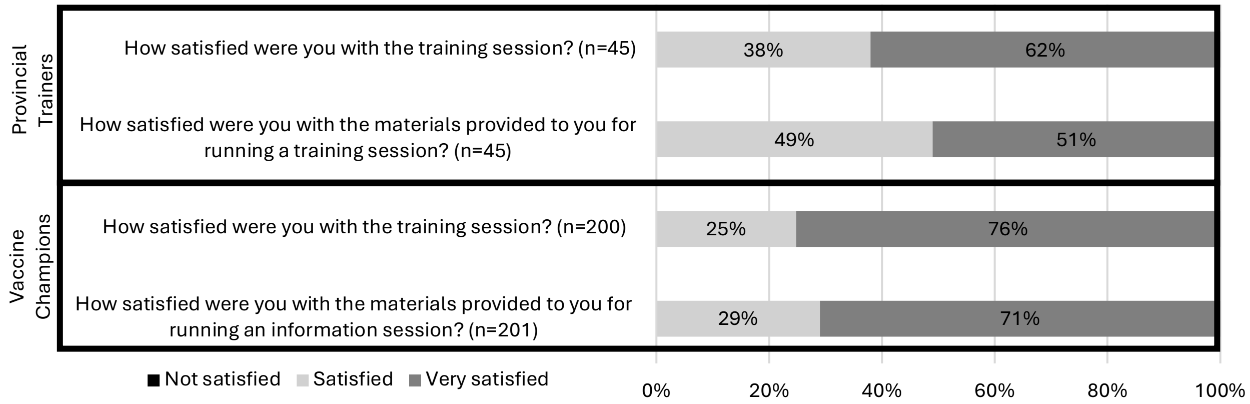


**Supplementary Figure 3: Provincial Trainer and Vaccine Champion satisfaction with training.** *Data were included for any participant who answered a post-training survey, regardless of whether they completed a pre-session survey.*
